# Causal mediation analysis of Subclinical Hypothyroidism in children with obesity and Non-Alcoholic Fatty Liver Disease

**DOI:** 10.1101/19004242

**Authors:** Presley Nichols, Yue Pan, Benjamin May, Martina Pavlicova, John Rausch, Ali Mencin, Vidhu V Thaker

## Abstract

**Background:** Nonalcoholic Fatty Liver Disease (NAFLD) is a common co-morbidity of obesity. Subclinical hypothyroidism (SH), also associated with obesity, may contribute to the dysmetabolic state that predisposes to NAFLD.

**Objective:** To assess the relationship between SH and NAFLD in children with biopsy-proven NAFLD compared to controls.

**Design and Methods:** In this retrospective study of children with biopsy-proven NAFLD and age-matched controls, the association of SH with NAFLD was assessed followed by causal mediation analyses under the counter-factual framework.

**Results:** Sixty-six cases and 4067 age-matched controls were included in the study. Children with NAFLD were more likely to be male (74.6 vs 39.4%, p < 0.001), have higher modified BMI-z scores (2.3 ±1.6 vs 1±1.6, p < 0.001), and abnormal metabolic parameters (TSH, ALT, HDL-C, non-HDL-C, LDL-C, and TG). Multivariate analyses controlling for age, sex and severity of obesity showed significant association between the 4^th^ quartile of TSH and NAFLD. Causal mediation analysis demonstrates that TSH mediates 44% of the effect of modified BMI-z score on NAFLD. This comprises of 16.2% (OR = 1.1, p < 0.001) caused by the indirect effect of TSH and its interaction with modified BMI-z, and 26.5% (OR = 1.1, p = 0.01) as an autonomous effect of TSH on NAFLD regardless of the obesity.

**Conclusions:** The association of SH and biopsy-proven NAFLD is demonstrated in children from predominantly Latino population. Further, a causal mediation analysis implicates an effect of TSH on NAFLD, independent of obesity.

## Introduction

Non-alcoholic fatty liver disease (NAFLD), defined as hepatic steatosis by imaging or histology without a secondary cause of hepatic fat accumulation^1^, has become the most common chronic liver disease in children in parallel with the rising prevalence of obesity ^2^. NAFLD is considered the hepatic manifestation of the metabolic syndrome which includes high blood pressure, dyslipidemia, insulin resistance and truncal obesity^3^. The spectrum of NAFLD includes simple steatosis through steatohepatitis. Simple steatosis may have a benign course, but steatohepatitis can lead to cirrhosis or hepatocellular carcinoma^4^. The gold standard for the diagnosis and staging of steatohepatitis is liver biopsy.

Subclinical hypothyroidism (SH) is defined as mildly elevated thyroid stimulating hormone (TSH 4.5-10 mIU/L) with normal levels of circulating thyroid hormone and commonly occurs with obesity in both pediatric and adult populations. Thyroid hormone is an important regulator of hepatic lipid metabolism through induction of genes involved in hepatic lipogenesis, coupling of autophagy to mitochondrial fat oxidation thereby leading to ketogenesis, and causing reverse cholesterol transport^5^. Therefore, it is not surprising that a close relationship has been seen between SH and cardiometabolic risk factors and NAFLD in adults^6-8^. A small number of studies in Caucasian children, primarily from Europe, have shown the association between SH and NAFLD in children and adolescents with obesity^9-12^. Using hepatic ultrasound for the diagnosis of NAFLD, these studies demonstrated higher levels of TSH in individuals with hepatic steatosis^9-12^.

The prevalence of NAFLD is higher in older males, especially those of Hispanic ethnicity^13,14^. However, little is known about the association of NAFLD and SH in this population. The goal of this study was to identify the relationship between SH and NAFLD in a cohort of children of predominantly Latino ancestry with biopsy-proven NAFLD. Further, given the role of thyroid hormones in lipid metabolism and NAFLD, we hypothesized that SH may be a causal mediator for the development of NAFLD in children with obesity.

## Materials and Methods

In this retrospective study, the cases were identified from a registry of children with biopsy-proven NAFLD from the Pediatric Gastroenterology and Hepatology program at Columbia University Irving Medical Center (CUIMC) between 2010-2018. The evaluation of the patients referred to the program includes investigation for infectious, autoimmune and heavy metal exposure as a cause of NAFLD in addition to the assessment of thyroid function. We selected the cases where etiology other than obesity was ruled out and data were available on thyroid function tests. All children underwent liver biopsy for persistently elevated serum aminotransferase level despite lifestyle interventions^1,4^ to confirm the diagnosis as well as to assess inflammation, fibrosis and the degree of hepatic fat deposition.

The control subjects were age-matched children attending the primary care clinics at the same institution with available TSH and free T4/total T4 levels, ALT ≤ 45 IU/L, AST ≤ 40 IU/L, bilirubin ≤ 1.5 mg/dl and alkaline phosphatase ≤150 IU/L, and absence of any acute or chronic liver disease seen in the same time period, 2010-2018. Retrospective lifetime data was collected from the electronic health records (EHRs) including prior physical examinations, anthropometric parameters measured during routine clinical care, laboratory tests, imaging studies and diagnoses codes. All subjects with ICD-codes consistent with renal disease, diabetes, known thyroid or other endocrine disease were excluded. Body mass index (BMI) was calculated as weight in kg/(height in m)^2^. BMI-z scores and modified BMI-z (mod-BMI-z) were calculated as per the CDC guidelines^15,16^. BMI class was considered normal BMI between 5-85^th^ percentile for age, moderate obesity as BMI between the 85^th^ percentile-120% of 95^th^ percentile, and severe obesity as BMI > 120% of 95^th^ percentile for age^17^. The population served by CUIMC is primarily children residing in the Washington Heights area of New York City with the racial/ethnic background of 71% Hispanic, 7% non-Hispanic Black, 17% non-Hispanic Whites, 3% Asian and 1% others according to the U.S. Census Bureau Population estimates, 2013, which is also the expected distribution of the population in this study. As the validity of individual level data for race and ethnicity could not be assured, this data was not included in the analyses.

### Liver Biopsy

Liver biopsy was performed in patients with evidence of hepatic steatosis on imaging with excluded secondary causes of liver disease and persistently elevated aminotransferases despite lifestyle interventions as per the standard of care guidelines^1,4^. Needle biopsy was performed in a pediatric procedure unit with anesthesia. Biopsy specimen was sent to the Columbia University pathology department for histological analysis of liver inflammation, fibrosis and fat deposition based on hematoxylin and eosin staining as well as trichrome staining when applicable. As per pathology guidelines, NAFLD was defined as macrovesicular steatosis in greater than 5% of hepatocytes^18^.

### Laboratory measurements

Results of laboratory tests were ascertained from the EHRs. Serum concentrations of total cholesterol (TC), high-density lipoprotein cholesterol (HDL-C) and triglycerides (TG) were measured using routine enzymatic methods with Roche/Hitachi Cobas® c system analyzer (USA). Value of low-density cholesterol (LDL-C) was calculated using Friedwald equation. Standard liver function tests, including ALT, AST, alkaline phosphatase and total bilirubin were measured on the same day with an auto analyzer. Non-HDL cholesterol was calculated as TC minus HDL-C. Serum samples were assayed for fT4, total T4 and TSH levels using an automated chemiluminescence assay system on Roche e601 platform. Peak values for TSH, ALT, TG, HDL-C, LDL-C and non-HDL cholesterol were extracted from individuals’ lab test results. Subjects were excluded if they have TSH value greater than 10 mIU/L or positive antithyroid antibodies and/or received thyroid related medications.

### Statistical Methods

A descriptive analysis of the cohort was performed by calculating absolute and relative proportions for categorical variables, and the mean and standard deviation for continuous variables. Distribution of continuous variables were examined for skewness and kurtosis and were logarithmically transformed, when appropriate. Geometric means and SD are reported where the variables were not normally distributed. To assess differences between the subjects with and without NAFLD, the Wilcoxon rank-sum test was applied for continuous variables and the chi-squared test for categorical variables. TSH values were categorized into four equal groups (quartiles) for the univariate and multivariate analysis. Univariate analysis was performed to assess the association between TSH quartiles and NAFLD. Multivariate logistic regression was used to determine the association between TSH quartiles and NAFLD adjusted for age, gender and class of obesity. Similar methods were also used for association between the individual lipid levels and TSH. For sensitivity analyses, the above analyses were also performed with TSH as a binary variable using a level > 4.5 mIU/L as abnormal.

To explore causal mediation of SH in the pathogenesis of NAFLD, mediation analysis using the counterfactual framework was performed between mod-BMI-z (continuous) with TSH as the mediator and presence of NAFLD (binary outcome) using logit link along with exposure mediator interaction ^19,20^. Mod-BMI-z was used as this has been shown to be a better measure of adiposity than BMI-z in children with severe obesity^15^. Age and gender were used as covariates in the causal mediation models. The mediation effect was decomposed into 4-components: i) direct effect : the effect of exposure, mod-BMI-z, on NAFLD in the absence of the mediator, TSH, ii) interactive effect : the effect of the mediator, TSH, on the outcome, NAFLD in the absence of the exposure, mod-BMI-z, iii) a mediated interaction: the interaction of exposure, mod-BMI-z and the mediator, TSH, and iv) pure mediated effect: mediation of TSH between the exposure and the outcome^21^. TSH and non-HDL-C levels were log-transformed and centered for these analyses. The analyses were performed in SAS v3.7 (SAS Institute, Cary, NC, USA) and R-statistical software v3.2 using libraries aov, dplyr, DescTools and ggplot2. A two tailed p-value < 0.05 was considered significant. The study protocol was approved by the Institutional Review Board at Columbia University Medical Center as an expedited exempt protocol due to the retrospective nature of the study.

## Results

From the registry of 78 children with biopsy-proven NAFLD, a total of 66 subjects with available thyroid studies were included as cases. There were no differences in the age, gender, BMI or lipoprotein profile of the excluded cases. Of the total 21,258 unique age-matched children seen at the primary care clinic in the same time period, 4300 subjects had data available on thyroid function tests and lipoproteins variables used in the study. One hundred eighty-three subjects were eliminated for elevated liver enzymes, and 50 for biologically implausible anthropometric values or chronic disease based on ICD-codes including thyroid disease. The final study cohort comprised of 66 cases with biopsy proven NAFLD and 4067 age matched controls. The demographic details of the cohort are provided in Table 1. Briefly, there were a higher number of males in the NAFLD group, the BMI z-score, mod-BMI-z and the proportion of individuals with severe obesity was higher in the NAFLD group compared to the controls. All the metabolic parameters assessed in the study (TSH, ALT, HDL-C, non-HDL-C, LDL-C and TG) were significantly higher in the cases with NAFLD, and free T4 levels were similar (Table 1). The high ALT levels of the cases are in keeping with the expectation of presence of Non-Alcoholic Steato-Hepatitis (NASH) in the cases^22^.

**Table 1.**
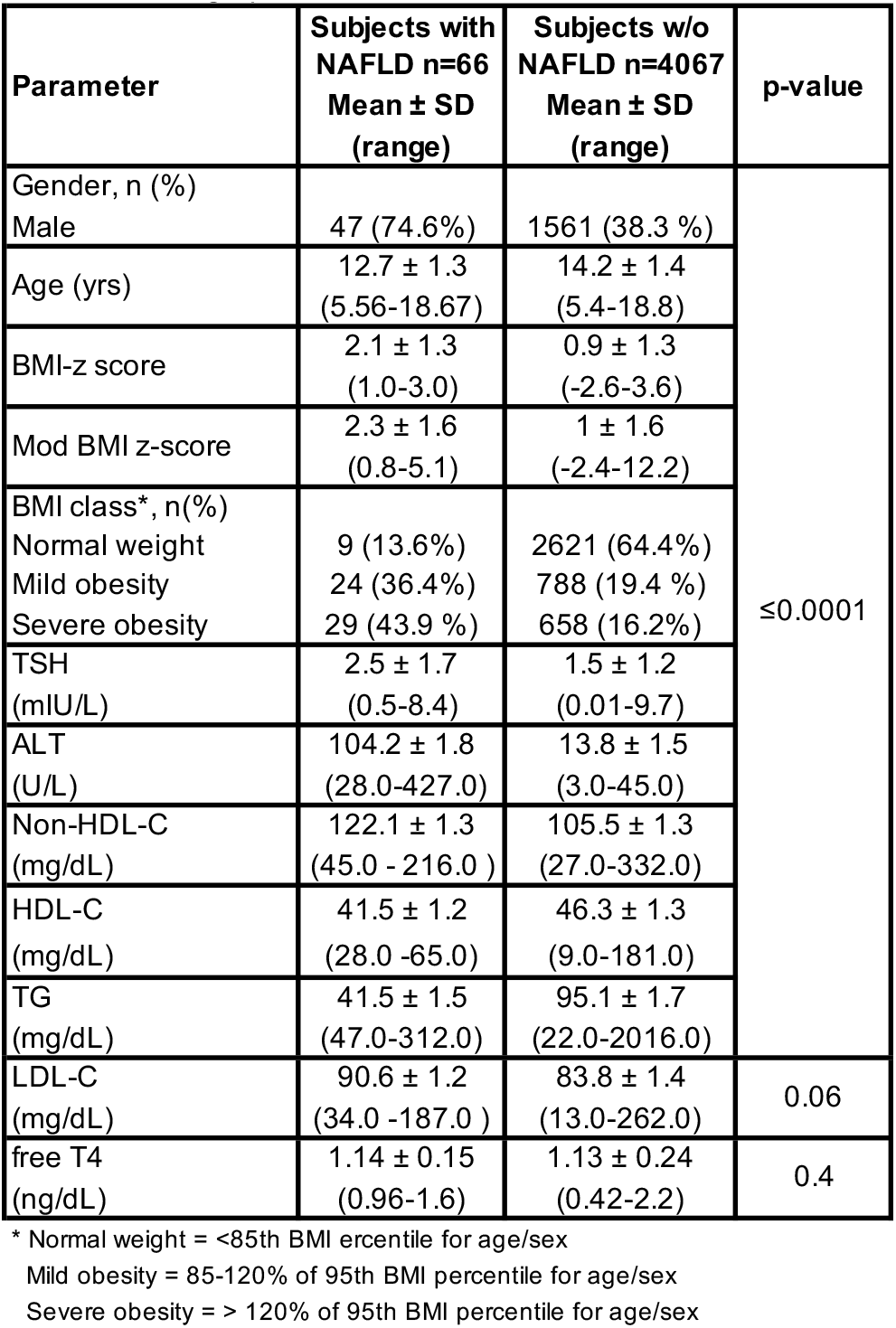
Demographic distribution of the cases and controls

| Parameter | Subjects with<br>NAFLD n=66<br>Mean ± SD<br>(range) | Subjects w/o<br>NAFLD n=4067<br>Mean ± SD<br>(range) | p-value |
| --- | --- | --- | --- |
| Gender, n (%) | | | $\leq 0.0001$ |
| Male | 47 (74.6%) | 1561 (38.3 %) |  |
| Age (yrs) | 12.7 ± 1.3<br>(5.56-18.67) | 14.2 ± 1.4<br>(5.4-18.8) | |
| BMI-z score | 2.1 ± 1.3<br>(1.0-3.0) | 0.9 ± 1.3<br>(-2.6-3.6) | |
| Mod BMI z-score | 2.3 ± 1.6<br>(0.8-5.1) | 1 ± 1.6<br>(-2.4-12.2) | |
| BMI class*, n(%) |  |  |  |
| Normal weight | 9 (13.6%) | 2621 (64.4%) |  |
| Mild obesity | 24 (36.4%) | 788 (19.4 %) |  |
| Severe obesity | 29 (43.9 %) | 658 (16.2%) |  |
| TSH<br>(mIU/L) | 2.5 ± 1.7<br>(0.5-8.4) | 1.5 ± 1.2<br>(0.01-9.7) | |
| ALT<br>(U/L) | 104.2 ± 1.8<br>(28.0-427.0) | 13.8 ± 1.5<br>(3.0-45.0) | |
| Non-HDL-C<br>(mg/dL) | 122.1 ± 1.3<br>(45.0 - 216.0 ) | 105.5 ± 1.3<br>(27.0-332.0) | |
| HDL-C<br>(mg/dL) | 41.5 ± 1.2<br>(28.0 -65.0) | 46.3 ± 1.3<br>(9.0-181.0) | |
| TG<br>(mg/dL) | 41.5 ± 1.5<br>(47.0-312.0) | 95.1 ± 1.7<br>(22.0-2016.0) | |
| LDL-C<br>(mg/dL) | 90.6 ± 1.2<br>(34.0 -187.0 ) | 83.8 ± 1.4<br>(13.0-262.0) | 0.06 |
| free T4<br>(ng/dL) | 1.14 ± 0.15<br>(0.96-1.6) | 1.13 ± 0.24<br>(0.42-2.2) | 0.4 |

\* Normal weight = <85th BMI ercentile for age/sex
Mild obesity = 85-120% of 95th BMI percentile for age/sex
Severe obesity = > 120% of 95th BMI percentile for age/sex

The TSH levels were divided into 4 quartiles: TSH Q1 < 1.06, TSH Q2 1.06-1.59, TSH Q3 1.59-2.35, TSH Q4 > 2.35 mU/L (Table 2). In the univariate analyses there were higher odds of the presence of NAFLD in the 3^rd^ and 4^th^ quartile of TSH compared to TSH Q1. Controlling for age, gender, and class of obesity, this association reached statistical significance in the 4^th^ quartile. ALT, non-HDL-C, and triglycerides showed a similar pattern but not LDL-C, HDL-C and TC (Table 2). While the total number of children across the 4 quartiles are similar (Table 2), the proportion of cases in the higher TSH quartiles is more compared to the controls (Fig. 1).

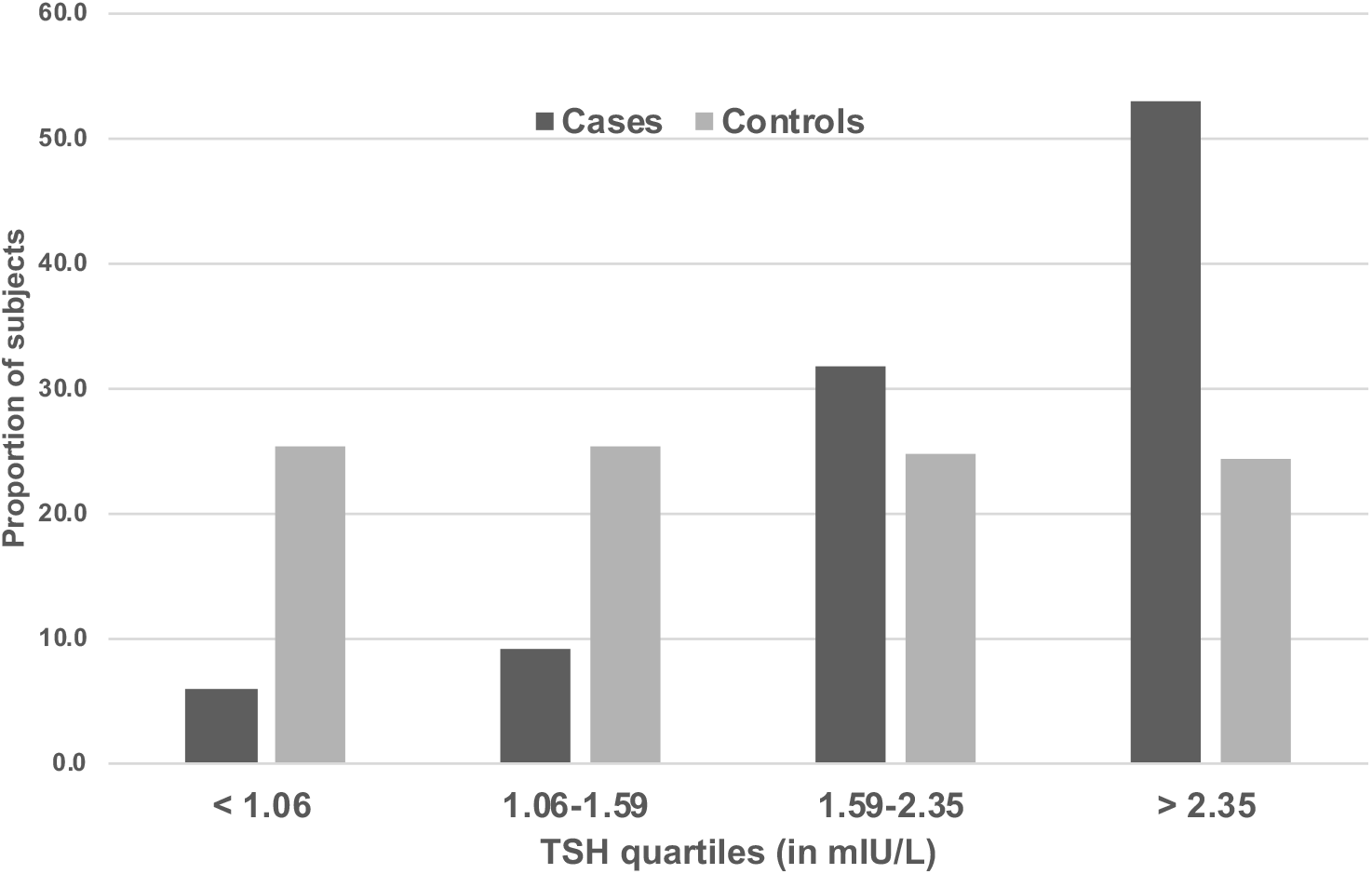

**Table 2.**
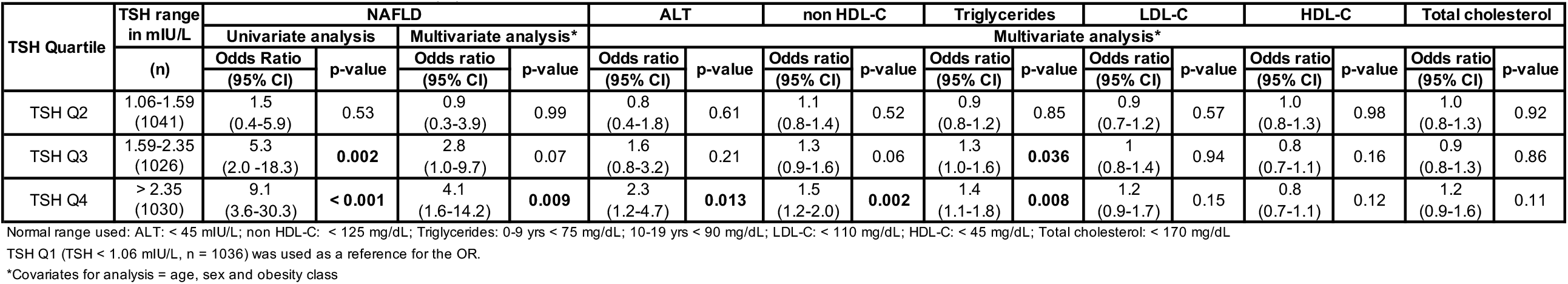
Association between TSH and NAFLD, ALT and lipoproteins

Sensitivity analyses with a TSH level of > 4.5 mIU/L considered as abnormal controlling for age, gender and BMI class showed a statistically significant association with NAFLD (OR = 3.7 [95% CI 1.6,7.5], p < 0.001) that persisted while controlling for non-HDL-C (OR = 3.5 [95% CI 1.5,7.2], p = 0.002). A similar association was not noted between abnormal TSH category and LDL-C, HDL-C, non-HDL-C, TC or TG.

The causal mediation analysis showed 57.3% (OR = 1.3 [95% CI 1.1,1.5], p = 0.001) direct effect of mod-BMI-z on the presence of NAFLD. The mediated effect of TSH on NAFLD was 16.17% (OR = 1.05 [95% CI 1.0-1.1], p < 0.001) comprising of 9.4% of pure mediation and 6.8% of mod-BMI-z and TSH interaction. Interestingly, 26.5% (OR = 1.13 [95% CI 1.0-1.3], p = 0.01) of the effect was the reference interaction caused by TSH on NAFLD, without the influence of mod-BMI-z (Table 3). The total interaction effect of TSH, both from its interaction with mod-BMI-z and the independent effect, was 33.3% (p = 0.002). Of the total effect of mod-BMI-z on NAFLD, 44% can be eliminated by intervention on the mediator, TSH.

**Table 3.**
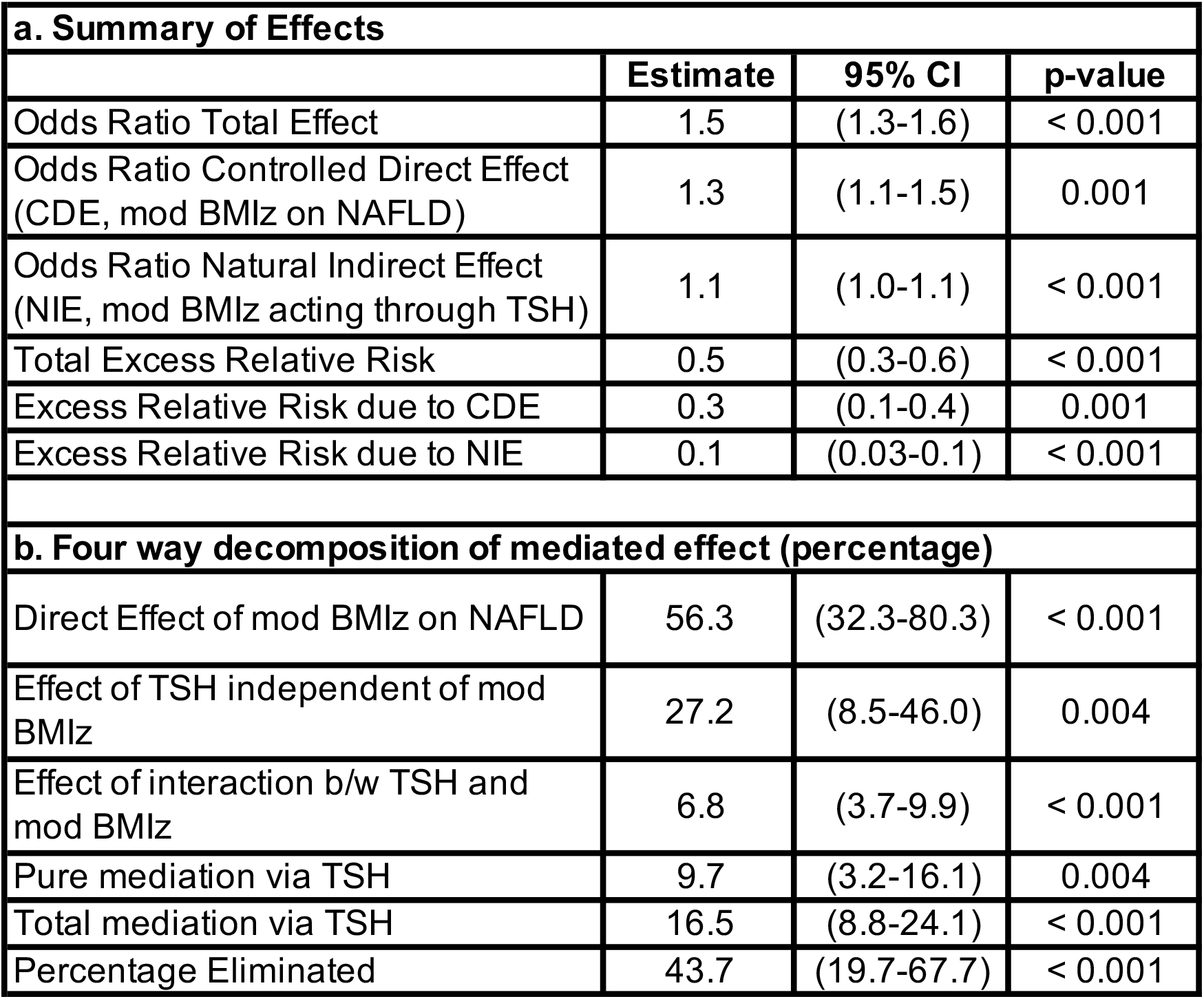
Causal Mediation Analysis of TSH as a mediator of mod BMIz and NAFLD.

| **a. Summary of Effects** |  |  |  |
| --- | --- | --- | --- |
|  | **Estimate** | **95% CI** | **p-value** |
| Odds Ratio Total Effect | 1.5 | (1.3-1.6) | < 0.001 |
| Odds Ratio Controlled Direct Effect (CDE, mod BMlz on NAFLD) | 1.3 | (1.1-1.5) | 0.001 |
| Odds Ratio Natural Indirect Effect (NIE, mod BMlz acting through TSH) | 1.1 | (1.0-1.1) | < 0.001 |
| Total Excess Relative Risk | 0.5 | (0.3-0.6) | < 0.001 |
| Excess Relative Risk due to CDE | 0.3 | (0.1-0.4) | 0.001 |
| Excess Relative Risk due to NIE | 0.1 | (0.03-0.1) | < 0.001 |
| **b. Four way decomposition of mediated effect (percentage)** |  |  |  |
| Direct Effect of mod BMlz on NAFLD | 56.3 | (32.3-80.3) | < 0.001 |
| Effect of TSH independent of mod BMlz | 27.2 | (8.5-46.0) | 0.004 |
| Effect of interaction b/w TSH and mod BMlz | 6.8 | (3.7-9.9) | < 0.001 |
| Pure mediation via TSH | 9.7 | (3.2-16.1) | 0.004 |
| Total mediation via TSH | 16.5 | (8.8-24.1) | < 0.001 |
| Percentage Eliminated | 43.7 | (19.7-67.7) | < 0.001 |

## Discussion

This study shows the association of SH and NAFLD in children of predominantly Latino ancestry with biopsy-proven diagnosis of NAFLD. To our knowledge, this is the first study in children to examine this relationship in cases with the gold-standard for diagnosis of NAFLD and explore the causal mediation of TSH for NAFLD. The causal mediation analysis demonstrates a partial mediation role of TSH in the relationship between the mod-BMI-z score and the presence of NAFLD. Additionally, the 4-way decomposition of the causal mediation model shows an effect of TSH on NAFLD independent of mod-BMI-z. Taken together these 2 effects provide statistical evidence for the mechanistic role of SH in the association between mod-BMI-z and NAFLD.

SH is a common occurrence in adults with a tendency towards development of overt hypothyroidism^23^. In adults, SH has been variably associated with adverse cardiometabolic effects, including dyslipidemia, insulin resistance, diastolic dysfunction, endothelial dysfunction, coronary heart disease and heart failure^6,24,25^. On the other hand, SH among children has been considered a benign and remitting condition, with the risk for progression only seen in individuals with certain underlying causes, such as autoimmune disease^26^. The evidence on the effect of SH on the growth and neurocognitive outcomes is conflicting^27,28^, but the evidence on the proatherogenic and metabolic abnormalities is increasingly being recognized^29-33^.

In this study, the children with NAFLD had a higher degree of obesity (BMI-z, mod-BMI-z and proportion of children with severe obesity) with higher levels of non-HDL-C and TG. This is similar to the published reports. In a retrospective cross-sectional analysis of 528 euthyroid children, Aypak *et al* found an association of higher levels of TSH and fT3 levels with obesity, as well as an association of increased level of dyslipidemia and higher systolic blood pressure with higher levels of TSH^29^. In a cohort of 49 prepubertal children with SH for a minimum of 2 years and age matched controls, Cerbone *et al* showed higher cardiometabolic risk factors, measured in the form of waist to height ratio, atherogenic index (ratio of TC: HDL-C), TG: HDL-C ratio, homocysteine levels, and lower HDL-C levels^30^. These findings were replicated in a cohort of adolescents ascertained from the community-based Korea National Health and Nutrition Examination surveys ^33^. Similar observations are made in a cohort of 22,147 children with Type 1 diabetes collected from multiple centers across Germany with or without the presence of autoantibodies^32^. Pacifico *et al* extended the observations of the association of metabolic findings with SH to NAFLD. Their study of 49 children with NAFLD and matched controls showed that in addition to the association with hypertriglyceridemia and insulin resistance, SH (TSH > 4.5 mIU/L) was also associated with NAFLD, while adjusting for age, gender, BMI and free T3/T4 levels^11^. In a separate study, higher insulin resistance, carotid intimal thickness and left ventricular mass was shown in children and adolescents with SH and NAFLD compared to matched obese and lean controls^34^.

As many of the human studies on SH have been performed in children with obesity, there is some debate about the directionality^28^. The inflammatory state in obesity is thought to influence the sodium-iodine symporter causing hyperthyrotropemia that may resolve with weight loss ^35^. It is not known whether hyperthyrotropinemia, dyslipidemia and NAFLD are driven by similar (or serial) mechanisms and in what direction. The mediation analysis performed in this study attempts to address this question. Given the known biology of the influence of thyroid hormone on lipid metabolism^5^, we hypothesized that TSH is a mediator of the effect of obesity on NAFLD. The causal mediation analysis under the counterfactual framework showed that the association between obesity and NAFLD is partially mediated by TSH. The more interesting finding from this analysis was the identification of the effect of TSH on NAFLD independent of the mod-BMI-z. This raises the question of whether children with SH in absence of obesity are also at a higher cardiometabolic risk. While studies of children with long-term untreated autoimmune and idiopathic SH do not show higher overall adiposity, they may suffer from higher visceral adiposity^36^. Indeed, waist circumference and waist-to-height ratio were higher among children with mild SH than among healthy euthyroid children matched for age, gender, height and pubertal status^36,37^.

Thyroid hormones have direct effects on both cholesterol and fatty acid synthesis and metabolism in a cell autonomous manner by transcriptional regulation of target genes involved in several of the hepatic metabolism pathways. Animal studies have shown that mildly hypothyroid mice develop NAFLD by impaired suppression of adipose tissue lipolysis as well as upregulation of de novo lipogenesis due to breakdown in the complex metabolic pathways in the liver^38^. Aside from this, TSH has been independently found to modulate lipid metabolism through the TSH receptors on the hepatocytes to induce hepatosteatosis via sterol regulatory element binding protein, SREBP^39^. TSH also suppresses the synthesis of hepatic bile acid via an SREBP2-hepatocyte nuclear factor 4α (HNFα)-CYP7A1 signaling pathway^40^. Moreover, TSH inhibits cholesterol synthesis by increasing AMPK-mediated phosphorylation of hydroxymethylglutaryl-CoA reductase (HMGCR) to inhibit HMGCR activity^41^. Collectively, these findings support the role of TSH itself in regulation of both hepatic lipid and cholesterol homeostasis.

The strengths of this study are the availability of the large cohort of children with NAFLD proven by liver biospy, and the matched control group that allowed the causal mediation analysis in a population of children of predominantly Latino ancestry. The limitations are the retrospective nature of the cohort, and data extraction from EHRs that precludes confirmation by repeating the tests or performing additional ones. We were unable to include measures such as race/ethnicity, glucose, insulin, blood pressure, waist circumference or pubertal status in this analysis because either the data were incomplete, or the accuracy of ascertainment could not be assured. Further, while we used the thyroid function tests obtained prior to the established diagnosis of NAFLD, it is possible that undetected NAFLD was already present at the time, in which case, the relationship is an association rather than mediation. Regardless, the results make an important contribution to the growing body of evidence on the complex relationship between thyroid hormone, lipid metabolism and its role in long-term cardiometabolic consequences of obesity, raising the question whether there is a role of treating SH, either by weight loss or medications, to improve the long-term health outcomes. The results of this study can be considered as the preliminary data to spark further conversation and longitudinal studies to understand these mechanisms in humans towards more effective clinical management strategies in future.

## Data Availability

The data used in the study is not freely available due to ethical restrictions and can be shared under appropriate IRB-approved data use agreements.

## Notes

### Competing Interest Statement

The authors have declared no competing interest.

### Clinical Trial

This is a retrospective study and not a clinical trial.

### Funding Statement

This work is supported in part by NIH-NIDDK K23 DK 110539 to VVT and the National Center for Advancing Translational Sciences, NIH, through Grant Number UL1TR001873.

### Author Declarations

All relevant ethical guidelines have been followed and any necessary IRB and/or ethics committee approvals have been obtained.

Any clinical trials involved have been registered with an ICMJE-approved registry such as ClinicalTrials.gov and the trial ID is included in the manuscript.

## References

1. Chalasani N, Younossi Z, Lavine JE, et al. The diagnosis and management of nonalcoholic fatty liver disease: Practice guidance from the American Association for the Study of Liver Diseases. Hepatology. 2018;67(1):328–357.

2. Anderson EL, Howe LD, Jones HE, Higgins JP, Lawlor DA, Fraser A. The Prevalence of Non-Alcoholic Fatty Liver Disease in Children and Adolescents: A Systematic Review and Meta-Analysis. PLoS One. 2015;10(10):e0140908.

3. Alterio A, Alisi A, Liccardo D, Nobili V. Non-alcoholic fatty liver and metabolic syndrome in children: a vicious circle. Horm Res Paediatr. 2014;82(5):283–289.

4. Vos MB, Abrams SH, Barlow SE, et al. NASPGHAN Clinical Practice Guideline for the Diagnosis and Treatment of Nonalcoholic Fatty Liver Disease in Children: Recommendations from the Expert Committee on NAFLD (ECON) and the North American Society of Pediatric Gastroenterology, Hepatology and Nutrition (NASPGHAN). J Pediatr Gastroenterol Nutr. 2017;64(2):319–334.

5. Sinha RA, Singh BK, Yen PM. Direct effects of thyroid hormones on hepatic lipid metabolism. Nat Rev Endocrinol. 2018;14(5):259–269.

6. Rodondi N, den Elzen WP, Bauer DC, et al. Subclinical hypothyroidism and the risk of coronary heart disease and mortality. Jama. 2010;304(12):1365–1374.

7. Guo Z, Li M, Han B, Qi X. Association of non-alcoholic fatty liver disease with thyroid function: A systematic review and meta-analysis. Digestive and liver disease : official journal of the Italian Society of Gastroenterology and the Italian Association for the Study of the Liver. 2018;50(11):1153–1162.

8. Gencer B, Collet TH, Virgini V, et al. Subclinical thyroid dysfunction and the risk of heart failure events: an individual participant data analysis from 6 prospective cohorts. Circulation. 2012;126(9):1040–1049.

9. Torun E, Ozgen IT, Gokce S, Aydin S, Cesur Y. Thyroid hormone levels in obese children and adolescents with non-alcoholic fatty liver disease. J Clin Res Pediatr Endocrinol. 2014;6(1):34–39.

10. Bilgin H, Pirgon O. Thyroid function in obese children with non-alcoholic fatty liver disease. J Clin Res Pediatr Endocrinol. 2014;6(3):152–157.

11. Pacifico L, Bonci E, Ferraro F, Andreoli G, Bascetta S, Chiesa C. Hepatic steatosis and thyroid function tests in overweight and obese children. International journal of endocrinology. 2013;2013:381014.

12. Kaltenbach TE, Graeter T, Oeztuerk S, et al. Thyroid dysfunction and hepatic steatosis in overweight children and adolescents. Pediatr Obes. 2017;12(1):67–74.

13. Nobili V, Alkhouri N, Alisi A, et al. Nonalcoholic fatty liver disease: a challenge for pediatricians. JAMA Pediatr. 2015;169(2):170–176.

14. Fernandes DM, Pantangi V, Azam M, et al. Pediatric Nonalcoholic Fatty Liver Disease in New York City: An Autopsy Study. J Pediatr. 2018;200:174–180.

15. Freedman DS, Butte NF, Taveras EM, et al. BMI z-Scores are a poor indicator of adiposity among 2- to 19-year-olds with very high BMIs, NHANES 1999-2000 to 2013- 2014. Obesity (Silver Spring). 2017;25(4):739–746.

16. CDC. A SAS Program for the 2000 CDC Growth Charts (ages 0 to < 20 years). 2016; https://www.cdc.gov/nccdphp/dnpao/growthcharts/resources/sas.htm. Accessed December 12, 2018.

17. Gulati AK, Kaplan DW, Daniels SR. Clinical tracking of severely obese children: a new growth chart. Pediatrics. 2012;130(6):1136–1140.

18. Brunt EM. Pathology of nonalcoholic fatty liver disease. Nature reviews Gastroenterology & hepatology. 2010;7(4):195–203.

19. Valeri L, Vanderweele TJ. Mediation analysis allowing for exposure-mediator interactions and causal interpretation: theoretical assumptions and implementation with SAS and SPSS macros. Psychological methods. 2013;18(2):137–150.

20. Valeri L, VanderWeele TJ. SAS macro for causal mediation analysis with survival data. Epidemiology. 2015;26(2):e23–24.

21. VanderWeele TJ. A unification of mediation and interaction: a 4-way decomposition. Epidemiology. 2014;25(5):749–761.

22. Schwimmer JB, Newton KP, Awai HI, et al. Paediatric gastroenterology evaluation of overweight and obese children referred from primary care for suspected non-alcoholic fatty liver disease. Alimentary pharmacology & therapeutics. 2013;38(10):1267–1277.

23. Cooper DS, Biondi B. Subclinical thyroid disease. Lancet (London, England). 2012;379(9821):1142–1154.

24. Gao N, Zhang W, Zhang YZ, Yang Q, Chen SH. Carotid intima-media thickness in patients with subclinical hypothyroidism: a meta-analysis. Atherosclerosis. 2013;227(1):18–25.

25. Razvi S, Weaver JU, Vanderpump MP, Pearce SH. The incidence of ischemic heart disease and mortality in people with subclinical hypothyroidism: reanalysis of the Whickham Survey cohort. J Clin Endocrinol Metab. 2010;95(4):1734–1740.

26. Monzani A, Prodam F, Rapa A, et al. Endocrine disorders in childhood and adolescence. Natural history of subclinical hypothyroidism in children and adolescents and potential effects of replacement therapy: a review. Eur J Endocrinol. 2013;168(1):R1–r11.

27. Cerbone M, Bravaccio C, Capalbo D, et al. Linear growth and intellectual outcome in children with long-term idiopathic subclinical hypothyroidism. Eur J Endocrinol. 2011;164(4):591–597.

28. Salerno M, Capalbo D, Cerbone M, De Luca F. Subclinical hypothyroidism in childhood - current knowledge and open issues. Nat Rev Endocrinol. 2016;12(12):734–746.

29. Aypak C, Turedi O, Yuce A, Gorpelioglu S. Thyroid-stimulating hormone (TSH) level in nutritionally obese children and metabolic co-morbidity. J Pediatr Endocrinol Metab. 2013;26(7-8):703–708.

30. Cerbone M, Capalbo D, Wasniewska M, et al. Cardiovascular risk factors in children with long-standing untreated idiopathic subclinical hypothyroidism. J Clin Endocrinol Metab. 2014;99(8):2697–2703.

31. Dahl M, Ohrt JD, Fonvig CE, et al. Subclinical Hypothyroidism in Danish Lean and Obese Children and Adolescents. J Clin Res Pediatr Endocrinol. 2017;9(1):8–16.

32. Denzer C, Karges B, Nake A, et al. Subclinical hypothyroidism and dyslipidemia in children and adolescents with type 1 diabetes mellitus. Eur J Endocrinol. 2013;168(4):601–608.

33. Lee MK, Kim YM, Sohn SY, Lee JH, Won YJ, Kim SH. Evaluation of the relationship of subclinical hypothyroidism with metabolic syndrome and its components in adolescents: a population-based study. Endocrine. 2019.

34. Sert A, Pirgon O, Aypar E, Yilmaz H, Odabas D. Subclinical hypothyroidism as a risk factor for the development of cardiovascular disease in obese adolescents with nonalcoholic fatty liver disease. Pediatr Cardiol. 2013;34(5):1166–1174.

35. Licenziati MR, Valerio G, Vetrani I, De Maria G, Liotta F, Radetti G. Altered Thyroid Function and Structure in Children and Adolescents Who Are Overweight and Obese: Reversal After Weight Loss. J Clin Endocrinol Metab. 2019;104(7):2757–2765.

36. Wasniewska M, Salerno M, Cassio A, et al. Prospective evaluation of the natural course of idiopathic subclinical hypothyroidism in childhood and adolescence. Eur J Endocrinol. 2009;160(3):417–421.

37. Radetti G, Gottardi E, Bona G, Corrias A, Salardi S, Loche S. The natural history of euthyroid Hashimoto’s thyroiditis in children. J Pediatr. 2006;149(6):827–832.

38. Ferrandino G, Kaspari RR, Spadaro O, et al. Pathogenesis of hypothyroidism-induced NAFLD is driven by intra- and extrahepatic mechanisms. Proc Natl Acad Sci U S A. 2017;114(43):E9172–e9180.

39. Yan F, Wang Q, Lu M, et al. Thyrotropin increases hepatic triglyceride content through upregulation of SREBP-1c activity. J Hepatol. 2014;61(6):1358–1364.

40. Song Y, Xu C, Shao S, et al. Thyroid-stimulating hormone regulates hepatic bile acid homeostasis via SREBP-2/HNF-4alpha/CYP7A1 axis. J Hepatol. 2015;62(5):1171–1179.

41. Zhang X, Song Y, Feng M, et al. Thyroid-stimulating hormone decreases HMG-CoA reductase phosphorylation via AMP-activated protein kinase in the liver. Journal of lipid research. 2015;56(5):963–971.

